# Youth susceptibility to tobacco use: Is it general or specific?

**DOI:** 10.1101/2021.05.25.21257808

**Authors:** Hui G. Cheng, Pavel N. Lizhnyak, Natasha A. Knight, Andrea R. Vansickel, Edward G. Largo

**Affiliations:** Altria Client Services LLC, 601 E. Jackson, Richmond VA 23219, USA

## Abstract

**Importance:** Susceptibility to tobacco use can help identify youth that are at risk for tobacco use.

**Objective:** To estimate the extent of overlap in susceptibilities across various tobacco products, investigate correlates with susceptibilities, and examine whether the relationship linking susceptibility with the onset of use is product specific or is accounted for by a general susceptibility-onset relationship.

**Design:** Prospective cohort study.

**Setting:** Analysis of data from the longitudinal Population Assessment of Tobacco and Health study wave 4 (December 2016 to January 2018) and wave 4.5 youth surveys (December 2017 to November 2018).

**Participants:** A nationally representative sample of non-institutionalized youth 12-17 years old who had never used a tobacco product at baseline assessment.

**Main variable of interest:** Susceptibility to the use of each type of tobacco product assessed at wave 4.

**Main outcomes:** Onset of use of various tobacco products defined as the first use occurring between waves 4 and 4.5 assessments.

**Results:** Cigarettes and e-cigarettes were the most common (∼25%), while snus was the least common (<5%), tobacco product to which youth were susceptible. There was a high degree of overlap in susceptibilities across tobacco products (65% of tobacco-susceptible youth were susceptible to more than one tobacco product). Tobacco-susceptible youth were more likely to have used cannabis or consumed alcohol in the past 30 days or to have tobacco-using peers. Susceptibility to use predicted the onset of use (incidence ratio = 3.2 to 12.9). Estimates for the product-specific path were null, except for e-cigarettes (β=0.08, 95% CI=0.04 to 0.13) and filtered cigars (β= -0.09, 95% CI= -0.13 to -0.05), after accounting for the general susceptibility-to-tobacco-onset relationship (β=0.50, 95% CI=0.42 to 0.58).

**Conclusions and Relevance:** Youth susceptibility to tobacco use overlaps widely across different tobacco products and other risky behaviors. Public health efforts may benefit from a holistic approach to risk behavior prevention planning.

**Key Points:** *Question:* Is susceptibility to tobacco use product specific, or does it represent a general openness to tobacco use?

*Findings:* There was a large degree of overlap in the susceptibility to tobacco use across various product categories. Associations linking susceptibility and the onset of use of a tobacco product were not statistically robust once the general level of susceptibility to tobacco use was considered.

*Meaning:* Susceptibility to tobacco use is better conceived as a general openness to tobacco use rather than product specific.

## 1. INTRODUCTION

The prevention of youth tobacco use is critical to public health. Susceptibility to tobacco use, defined as the lack of a determined decision not to use tobacco, is a robust predictor of youth tobacco initiation.^1^ Understanding the susceptibility to tobacco use helps identify adolescents who are at high risk for tobacco use and can aid the design of prevention strategies. Numerous studies have shown that youth who were susceptible to a tobacco product were more likely to start using that tobacco product. For example, Pierce and colleagues found that youth never smokers who were susceptible to smoking were five times as likely to start smoking in the subsequent four years compared to those who were not susceptible.^1^ In the past decade, e-cigarettes have become the most commonly used tobacco product among youth in the United States (US).^2,3^ Several recent studies have found that youth who were susceptible to e-cigarette use at baseline were more likely to start using e-cigarettes during follow-up.^4,5^ These studies provided product-specific evidence for the susceptibility-use relationship.

Recently, mounting evidence has suggested that the susceptibility-use relationship may not be product specific. For example, in a longitudinal study of Canadian 9^th^-12^th^ graders, susceptibility to cigarette smoking at baseline predicted the use of a wide range of tobacco products, including e-cigarette, cigarillo, and little cigar in addition to cigarette smoking.^6^ Similar findings have been documented in the US.^7^ Focusing on e-cigarettes and cigarettes, Nicksic and Barnes (2019) found evidence supporting cross-product prediction. That is, susceptibility to either e-cigarette use or cigarette smoking predicted the initiation of cigarette smoking and the initiation of e-cigarette use. Further inspection revealed a large degree of overlap in the susceptibility to cigarette smoking and the susceptibility to e-cigarette use (i.e., 67% of youth who were susceptible to cigarette smoking were also susceptible to e-cigarette use).^8^ It is well known that youth tobacco use is characterized by high levels of concurrent use of multiple tobacco products.^3,9^ In a previous study, we found that youth use of various tobacco products may be manifestations of a tendency to use tobacco in general.^10^ In this context, it is of interest to query whether the susceptibility-use relationship operates in a product-specific manner or reflects a general tendency towards tobacco use.

In the current study, we set out to investigate whether susceptibility to tobacco use is product specific or represents a general tendency towards multiple tobacco product use with the following specific aims: (a) to systematically estimate youth susceptibilities to various tobacco products and the degree of overlap; (b) to estimate associations between selected sociodemographic and behavioral variables and susceptibilities to various tobacco products; and (c) to estimate prospective relationships between susceptibility and the onset of various tobacco products among youth 12-17 years of age living in the US using recent data from a nationally representative longitudinal study. In this study, we used a structural equation modeling approach to estimate product-specific prediction while accounting for a general level of susceptibility.

## 2. METHODS

### 2.1 Study Population

In this study, the population of interest is US non-institutionalized civilian adolescents 12-17 years of age who had never used a tobacco product. Data were from the longitudinal Population Assessment of Tobacco and Health (PATH) study wave 4 (Dec. 2016-Jan. 2018) and wave 4.5 (Dec. 2017-Nov. 2018) surveys, which used a multi-stage sampling method to draw nationally representative samples after Institutional-Review-Board-approved parent consent and youth assent.^11^ In contrast to school surveys of adolescents, the PATH sample includes young people irrespective of school attendance, and its sampling frame includes college dormitories and children of active-duty military living in the US. More details about the PATH methodology is provided elsewhere.^11^ PATH Public Use data files were downloaded from https://www.icpsr.umich.edu/icpsrweb/NAHDAP/studies/36498 on Dec. 13, 2019.

### 2.2 Assessment

Audio computer assisted self-interviews (ACASI) with standardized multi-item modules were used to assess tobacco use history and a range of related variables. Never tobacco users at wave 4 were individuals who had never used any of the tobacco products assessed, even one time. Products assessed included cigarettes, e-cigarettes, cigars (traditional cigars, cigarillos, and filtered cigars), smokeless tobacco, snus, hookah, pipe, dissolvable tobacco, bidis, and kretek. Survey questions about ever use of these tobacco products were typically in the format of “Have you ever smoked/used …, even one or two puffs/times?”

Susceptibility to tobacco use was assessed via the following questions:

▪ “Have you ever been curious about using/smoking …?”
▪ “Do you think you will try/smoke … in the next year?”
▪ “Do you think that you will try/smoke … soon?”
▪ “If one of your best friends were to offer you a …, would you try/smoke it?”

Sets of these susceptibility questions were asked for cigarettes, cigars, cigarillos, filtered cigars, e-cigarettes, hookah, snus, and smokeless tobacco. Each question was rated on a 4-point Likert scale. In line with the literature, we dichotomized susceptibility into non-susceptible and susceptible.^1,8,12^ The former group included youth who answered “not at all curious” to the first question and “definitely not” to the other three questions. Any non “definitely not/not at all” answer to any of these four questions qualified the youth as susceptible to use of the tobacco product assessed. These measures were adapted from validated items of susceptibility to cigarette smoking.^1,12,13^

Incident use of a tobacco product was defined as using the product for the first time between wave 4 and wave 4.5 assessments among never users at wave 4.

Information about sex (male or female), age categories (12-14 or 15-17 years of age at baseline), and race/ethnicity is from survey items in the Demographics module. When these items were missing, information from the household screening roster was drawn. Other covariates of interest included cannabis use during the past 30 days, alcohol drinking during the past 30 days, peer tobacco use, school performance during the past 12 months (dichotomized as ‘mostly A’s and B’s’), and availability of tobacco at home. Cannabis use included using cannabis (marijuana, hash, THC, grass, pot, or weed) as well as blunts. Peer tobacco use was assessed via questions worded “how many of your best friends use/smoke …?” Separate questions were asked for cigarettes, e-cigarettes, traditional cigars, cigarillos, filtered cigars, snus, and smokeless tobacco. In this study, we dichotomized peer use into “none” and “any” for each tobacco product category assessed. Cannabis use, alcohol drinking, and peer tobacco use were based on the adolescent’s self-report. School performance and availability of tobacco products at home were based on information about the adolescent provided by a parent/guardian. Supplemental Table S1 provides details about the assessment of these covariates.

### 2.3 Analysis

First, we estimated the proportion of adolescent never tobacco users who were susceptible to each of the tobacco products. We then used a Venn diagram to visualize the degree of overlap in susceptibility to various tobacco products. To ease the visualization, we categorized tobacco products into three groups – combusted tobacco products (i.e., cigarettes, cigars, hookah), e-cigarettes, and oral tobacco products (i.e., smokeless tobacco and snus).

Next, we estimated the susceptibility to tobacco products by sociodemographic characteristics and behavioral variables.

To assess whether wave 4 susceptibility predicted the onset of tobacco product use, we used generalized linear regression with a log link to estimate incidence ratios for each tobacco product category studied here. In the last analysis step, we employed a structural equation modeling approach to estimate the product-specific susceptibility-use relationship while accounting for a general susceptibility-use relationship, as depicted in Figure 1. Measurement models were evaluated before the structural paths were drawn. Multiple fit indices were used to evaluate the goodness of fit, including root mean square of approximation (RMSEA),^14^ comparative fit index (CFI),^15^ and Tucker-Lewis index (TLI). A RMSEA<0.08 and CFI/TLI>0.90 were considered as indications of reasonably good model fit.^16,17^

**Figure 1.**
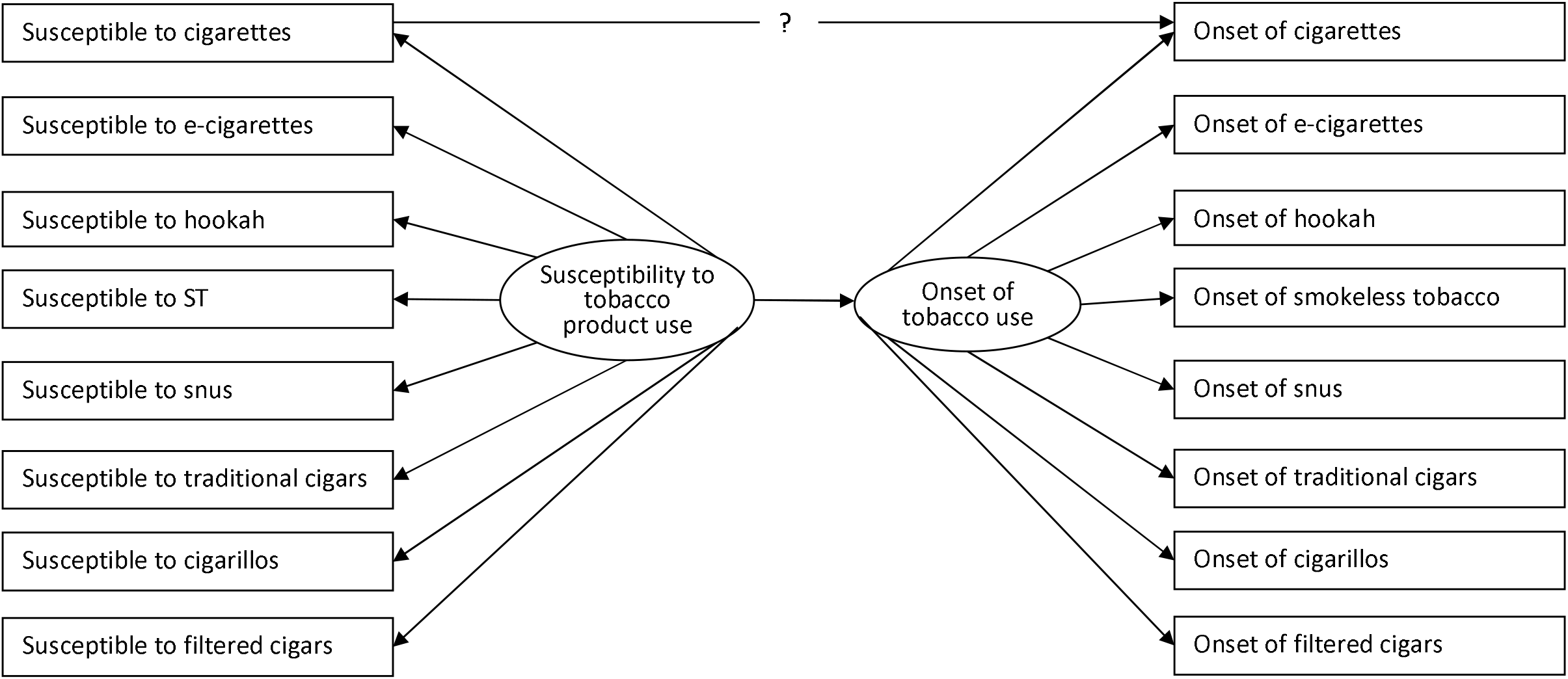
Depiction of a conceptual structural equation model to predict the onset of first cigarette smoking by susceptibility to smoking adjusting for a latent ‘susceptibility to use tobacco’ construct.

All analyses were weighted. Wave 4 cross-sectional weights were used for wave 4 cross-sectional analysis. Wave 4.5 longitudinal weights for wave 4 cohort were used for wave 4 and 4.5 prospective analysis. These weights incorporate adjustment for selection probability, nonresponse patterns, possible deficiencies in the sampling frame, and attrition.^11^ Variances of estimates were produced using balance repeat replication methods (Fay’s method with fay=0.3). A robust weighted least square mean and variance (WLSMV) adjusted estimator, which uses a full weight matrix, was used to accommodate categorical variables and complex survey design in structural equation models. Analyses were conducted using Stata 16.0 (StataCorp, College Station, Texas, USA) and Mplus 8.1 (Muthén & Muthén, Los Angeles, CA, USA).

## 3. RESULTS

### 3.1 Susceptibility to tobacco products among never users

At wave 4, there were a total of 10,976 never tobacco users, of whom 50% were girls, 59% were 12-14 years of age, 52% were non-Hispanic White, 14% were non-Hispanic Black, 23% were Hispanics, and 10% were other race/ethnicity groups. Most youth never tobacco users (63%) were not susceptible to any tobacco use. (For all susceptibility measures, there were <1% missing values.) As shown in Figure 2, cigarettes and e-cigarettes were the most common tobacco products that youth were susceptible to, and snus was the least common with less than 5% of youth never users susceptible to snus use.

**Figure 2.**
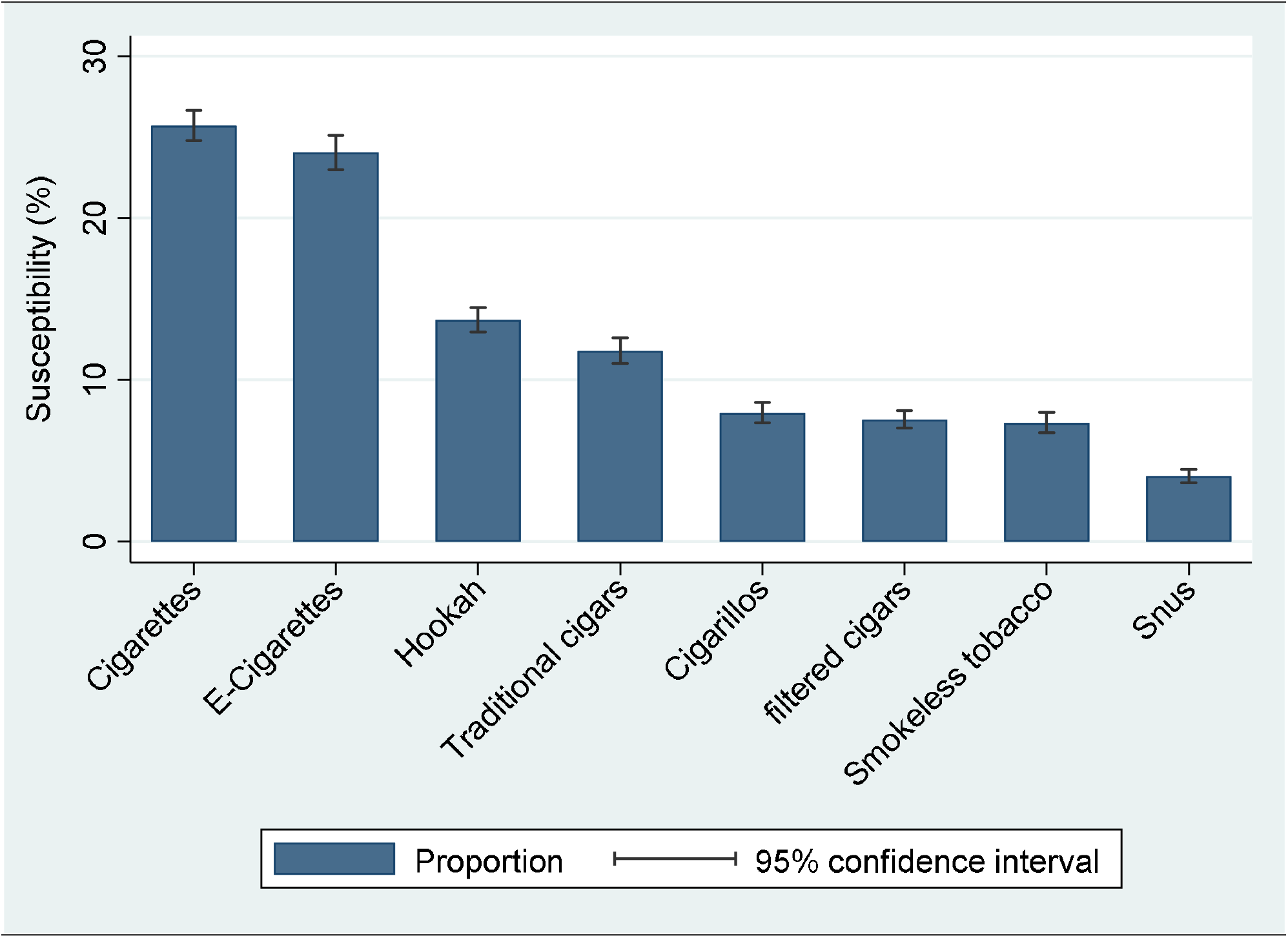
Estimated susceptibility to various tobacco products among youth never tobacco users. Data from PATH wave 4 youth survey.

The Venn diagram revealed a large degree of overlap in susceptibilities to the three tobacco product categories (see Figure 3). Fifty-eight percent of youth never users who were susceptible to any tobacco use were susceptible to at least two categories. When examined at the individual product level, 65% of youth never users who were susceptible to any tobacco use were susceptible to more than one product.

**Figure 3.**
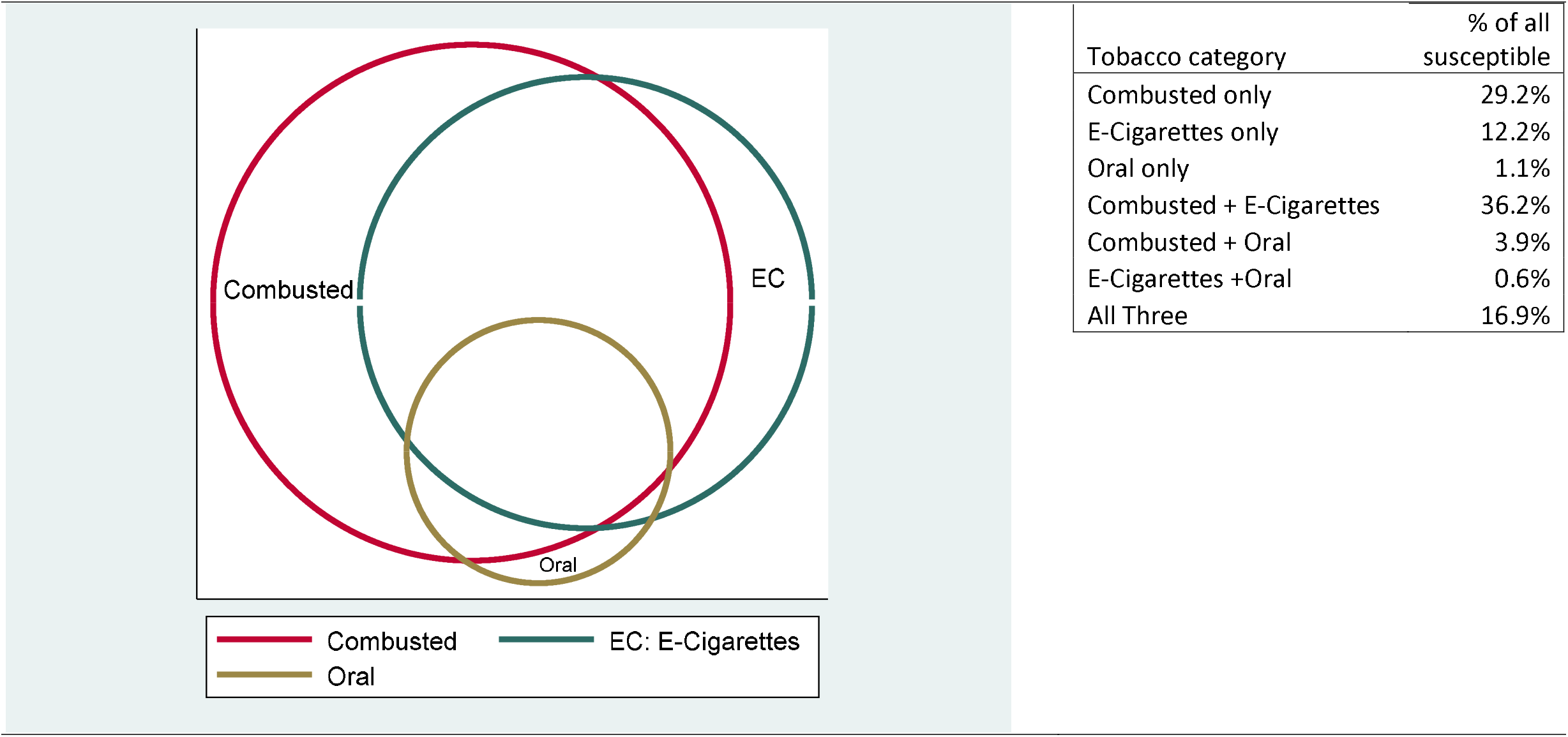
Relations of susceptibility to combusted tobacco products (cigarettes, cigars, and hookah), e-cigarettes, and oral tobacco products (snus and smokeless). Data from PATH wave 4 youth survey.

### 3.2 Characteristics of youth who are susceptible to each and any tobacco product

With respect to demographic characteristics, older adolescents (15-17 years of age) were more likely to be susceptible to all tobacco products assessed compared to younger adolescents (12-14 years of age, Table 1). With respect to behavioral and environmental variables assessed, adolescents who were current cannabis users and/or alcohol drinkers were more likely to be susceptible to tobacco use (Table 1). In addition, those who affiliated with peers who used tobacco were more likely to be susceptible to use tobacco (Table 1).

**Table 1.**
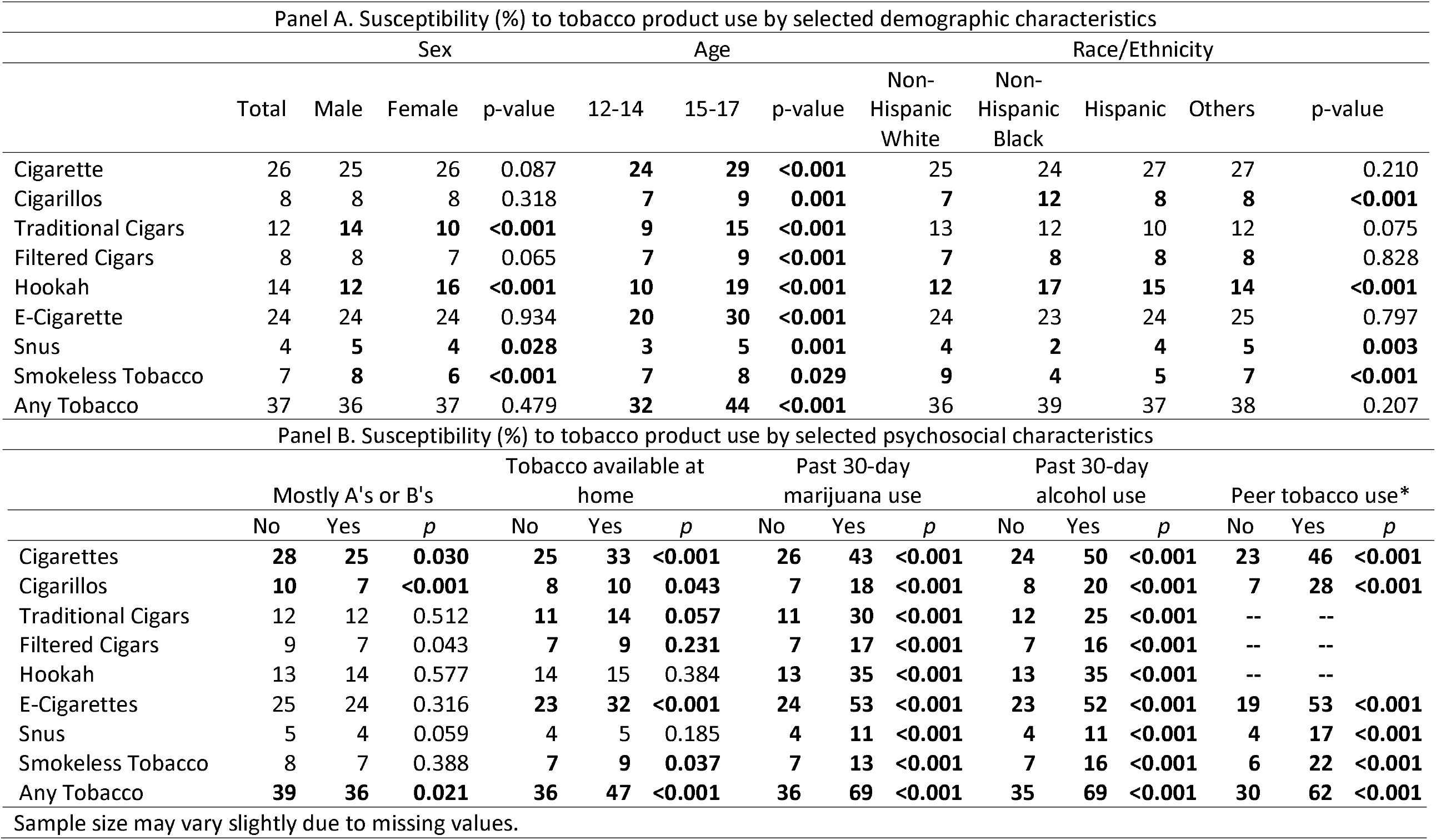
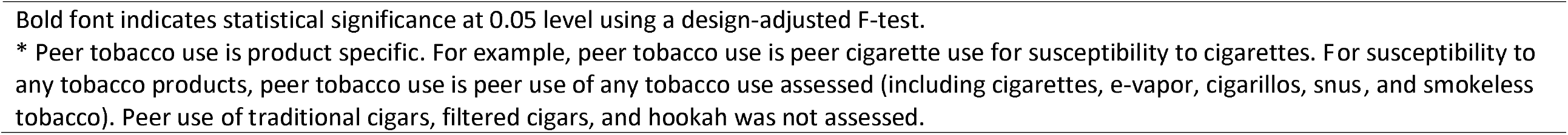
Susceptibility (%) to tobacco product use by selected demographic characteristics among youth never tobacco users. Data from PATH wave 4 youth survey 2017-2018 (n=10,977)

| Panel A. Susceptibility (%) to tobacco product use by selected demographic characteristics |  |  |  |  |  |  |  |  |  |  |  |  |  |  |  |
| --- | --- | --- | --- | --- | --- | --- | --- | --- | --- | --- | --- | --- | --- | --- | --- |
|  | Sex |  |  |  | Age |  |  | Race/Ethnicity |  |  |  |  |  |  |  |
|  | Total | Male | Female | p-value | 12-14 | 15-17 | p-value | Non-Hispanic White | Non-Hispanic Black | Hispanic | Others | p-value |  |  |  |
| Cigarette | 26 | 25 | 26 | 0.087 | 24 | 29 | <0.001 | 25 | 24 | 27 | 27 | 0.210 |  |  |  |
| Cigarillos | 8 | 8 | 8 | 0.318 | 7 | 9 | 0.001 | 7 | 12 | 8 | 8 | <0.001 |  |  |  |
| Traditional Cigars | 12 | 14 | 10 | <0.001 | 9 | 15 | <0.001 | 13 | 12 | 10 | 12 | 0.075 |  |  |  |
| Filtered Cigars | 8 | 8 | 7 | 0.065 | 7 | 9 | <0.001 | 7 | 8 | 8 | 8 | 0.828 |  |  |  |
| Hookah | 14 | 12 | 16 | <0.001 | 10 | 19 | <0.001 | 12 | 17 | 15 | 14 | <0.001 |  |  |  |
| E-Cigarette | 24 | 24 | 24 | 0.934 | 20 | 30 | <0.001 | 24 | 23 | 24 | 25 | 0.797 |  |  |  |
| Snus | 4 | 5 | 4 | 0.028 | 3 | 5 | 0.001 | 4 | 2 | 4 | 5 | 0.003 |  |  |  |
| Smokeless Tobacco | 7 | 8 | 6 | <0.001 | 7 | 8 | 0.029 | 9 | 4 | 5 | 7 | <0.001 |  |  |  |
| Any Tobacco | 37 | 36 | 37 | 0.479 | 32 | 44 | <0.001 | 36 | 39 | 37 | 38 | 0.207 |  |  |  |
| Panel B. Susceptibility (%) to tobacco product use by selected psychosocial characteristics |  |  |  |  |  |  |  |  |  |  |  |  |  |  |  |
|  | Mostly A's or B's |  |  | Tobacco available at home |  |  | Past 30-day marijuana use |  |  | Past 30-day alcohol use |  |  | Peer tobacco use* |  |  |
|  | No | Yes | p | No | Yes | p | No | Yes | p | No | Yes | p | No | Yes | p |
| Cigarettes | 28 | 25 | 0.030 | 25 | 33 | <0.001 | 26 | 43 | <0.001 | 24 | 50 | <0.001 | 23 | 46 | <0.001 |
| Cigarillos | 10 | 7 | <0.001 | 8 | 10 | 0.043 | 7 | 18 | <0.001 | 8 | 20 | <0.001 | 7 | 28 | <0.001 |
| Traditional Cigars | 12 | 12 | 0.512 | 11 | 14 | 0.057 | 11 | 30 | <0.001 | 12 | 25 | <0.001 | -- | -- |  |
| Filtered Cigars | 9 | 7 | 0.043 | 7 | 9 | 0.231 | 7 | 17 | <0.001 | 7 | 16 | <0.001 | -- | -- |  |
| Hookah | 13 | 14 | 0.577 | 14 | 15 | 0.384 | 13 | 35 | <0.001 | 13 | 35 | <0.001 | -- | -- |  |
| E-Cigarettes | 25 | 24 | 0.316 | 23 | 32 | <0.001 | 24 | 53 | <0.001 | 23 | 52 | <0.001 | 19 | 53 | <0.001 |
| Snus | 5 | 4 | 0.059 | 4 | 5 | 0.185 | 4 | 11 | <0.001 | 4 | 11 | <0.001 | 4 | 17 | <0.001 |
| Smokeless Tobacco | 8 | 7 | 0.388 | 7 | 9 | 0.037 | 7 | 13 | <0.001 | 7 | 16 | <0.001 | 6 | 22 | <0.001 |
| Any Tobacco | 39 | 36 | 0.021 | 36 | 47 | <0.001 | 36 | 69 | <0.001 | 35 | 69 | <0.001 | 30 | 62 | <0.001 |
| Sample size may vary slightly due to missing values. |  |  |  |  |  |  |  |  |  |  |  |  |  |  |  |

Bold font indicates statistical significance at 0.05 level using a design-adjusted F-test.
\* Peer tobacco use is product specific. For example, peer tobacco use is peer cigarette use for susceptibility to cigarettes. For susceptibility to any tobacco products, peer tobacco use is peer use of any tobacco use assessed (including cigarettes, e-vapor, cigarillos, snus, and smokeless tobacco). Peer use of traditional cigars, filtered cigars, and hookah was not assessed.

### 3.3 Prediction of susceptibility of the onset of tobacco product use

#### 3.3.1 Bivariate product-specific prediction

The onset of tobacco use was below 5%, irrespective of whether adolescents were susceptible or not for all tobacco products, except for e-cigarettes (Table 2). Adolescents who were susceptible to a tobacco product were more likely to use the product for the first time within the next year (except for filtered cigars, for which the association was not statistically significant). For example, 18.5% of adolescent never users who were susceptible to e-cigarettes at wave 4 assessment used e-cigarettes for the first time by the time of the wave 4.5 assessment, whereas 4.3% of those who were *not* susceptible to e-cigarettes did so (incidence ratio=4.3, 95% CI=3.6 to 5.0; Table 2).

**Table 2.** Onset of tobacco product use (%) stratified by susceptibility among youth never tobacco users, estimated incidence ratio, and estimates of the product-specific path for susceptibility predicting onset from structural equation models among youth never tobacco users. Data from PATH wave 4 and 4.5 youth surveys (n=8841).

|  | Not susceptible | Susceptible | IR<sup>1</sup> (95% CI) | Beta<sup>2</sup> (95% CI) |
| --- | --- | --- | --- | --- |
| Cigarettes | 1.1 | 3.6 | **3.2 (2.3, 4.6)** | -0.04 (-0.13, 0.05) |
| Cigars | 0.1 | 0.9 | **6.7 (2.5, 18.4)** | **0.08 (0.04, 0.13)** |
| Cigarillos | 0.4 | 4.2 | **9.4 (5.6, 16.0)** | 0.01 (-0.15, 0.17) |
| Filtered cigars | 0.1 | 0.3 | 3.8 (0.5, 27.9) | 0.06 (-0.01, 0.13) |
| Hookah | 0.4 | 1.6 | **3.9 (1.9, 8.1)** | **-0.09 (-0.13, -0.05)** |
| E-Cigarettes | 4.3 | 18.5 | **4.3 (3.6, 5.0)** | -0.03 (-0.15, 0.09) |
| Snus | 0.2 | 1.6 | **7.6 (2.4, 24.3)** | -0.02 (-0.17, 0.14) |
| Smokeless Tobacco | 0.2 | 2.6 | **12.9 (6.4, 26.1)** | 0.01 (-0.11, 0.12) |

<sup>1</sup>IR: incidence ratio estimated using generalized linear regression with a log link.
<sup>2</sup>Beta coefficients for the product-specific path were from the structural equation model, in which the estimate linking the susceptibility latent construct and the tobacco onset latent construct was 0.50 (95% CI = 0.42 to 0.58).
Bold font indicates statistical significance at 0.05 level.

#### 3.3.2 Results from the structural equation model

Due to the large overlap in susceptibility across different tobacco products, structural equation models were built to estimate the relationship of wave 4 susceptibility to a specific tobacco product and wave 4.5 onset of the tobacco product use accounting for the general level of susceptibility to various tobacco products and the general level of tobacco use onset (Figure 1). The latent construct for susceptibility represents the level of openness to tobacco use, and the latent construct for tobacco use onset represents the level of liability to starting tobacco use. In this model, the estimate of the direct path leading from the susceptibility to cigarette smoking to the onset of cigarette smoking presents a cigarette-specific susceptibility-onset relationship after accounting for the general level of susceptibility to tobacco use and the general level of tobacco onset.

Confirmatory factor analysis showed reasonable goodness of fit of a one-factor model that a single construct of susceptibility gives rise to the susceptibility to various tobacco products (RMSEA=0.052, CFI=0. 991, and TLI=0.988), as well as the unidimensionality of the onset of tobacco use (RMSEA=0.019, CFI=0. 950, and TLI=0.931) among wave 4 never tobacco users who were followed up at wave 4.5 (n=8841).

Next, a series of structural equation models, as depicted in Figure 1, were fit to the data to estimate the path for each specific tobacco product. These models fit the data reasonably well (RMSEA<0.05, CFI>0. 90, and TLI>0.90). The susceptibility latent construct was a robust predictor of the latent tobacco onset construct (β=0.50, 95% CI=0.42 to 0.58). Estimates of the product-specific path were not statistically significant after accounting for the path between general susceptibility and general tobacco onset (Table 2), except for e-cigarettes (positive prediction: β=0.08, 95% CI=0.04 to 0.13) and filtered cigars (inverse prediction: β= -0.09, 95% CI= -0.13 to -0.05). Nonetheless, the magnitude of these product-specific estimates was much smaller compared to the estimate linking the two latent constructs.

## 4. DISCUSSION

Results of these analyses showed that youth susceptibility to tobacco product use typically presents as a general openness to tobacco use rather than product-specific susceptibility. First, we found substantial overlap in susceptibility across different tobacco products. Second, this general susceptibility to tobacco use coincides with other risk-taking behaviors, such as current users of alcohol and cannabis and socializing with peers who use tobacco. Lastly, the prediction linking susceptibility to tobacco use onset operates primarily at the general tobacco susceptibility level. These findings indicate that a holistic approach towards adverse youth risk behaviors may be more effective in identifying youth who are at high-risk for tobacco use compared to any product-specific approaches.

The substantial overlap in susceptibility to tobacco use observed in this study extended previous evidence on e-cigarette and cigarettes susceptibility^8^ by including various forms of tobacco products and found that the majority (65%) of youth susceptible to using one tobacco product were susceptible to using multiple tobacco products. In addition, youth who used alcohol or cannabis and those who affiliated with tobacco-using peers, were more likely to be susceptible to tobacco use. These findings suggest that risky behaviors, including the use of other psychoactive substances and peer affiliation, can serve as indicators for youth at high risk for tobacco use. This is in line with previous studies showing cross-category prediction - youth never users who were susceptible to tobacco use were more likely to start using tobacco, alcohol, and other psychoactive drugs compared to youth who were not susceptible to tobacco use.^8,18^ The Common Liability Theory may best-explain risky behaviors as it accounts for socio-cultural, structural, and heritable traits, emphasizing individual liability rather than substance/product-specific attributes as contributing to substance use.^19^ Some recent studies have highlighted the role of an underlying liability towards tobacco use.^10,20^ The current study extended the previous studies on tobacco use behavior to the susceptibility to tobacco use.

Historically, males and individuals with poorer school performance have been found to be more likely to use or be susceptible to tobacco.^1,8,21,22^ In this study, we found that the association between tobacco susceptibility and sex, race/ethnicity, and school performance was not consistently present for all tobacco products, suggesting these demographic differences may change with the type of tobacco product. These findings align with a previous study on tobacco use (Wang et al., 2019) showing that demographic differences varied across tobacco product categories among youth tobacco users.^9^ It is important for teachers, parents, and public health professionals to consider these variations in their tobacco prevention work as the youth tobacco use landscape can change rapidly. In the meantime, use of other psychoactive drugs and peer affiliation may better indicate youth tobacco susceptibility.

Building upon previous findings on product-specific and cross-product prediction,^4,6-8^ we found that shared susceptibility is a key predictor of the onset of tobacco use. That is, even though product-specific predictions were strong when studied bivariately, they became relatively small and not statistically robust once the link between general susceptibility and general tobacco onset was considered. In this study, we observed a weak but statistically robust product-specific path for e-cigarettes suggesting that there is an e-cigarette-specific prediction that is not completely accounted for by the general susceptibility-onset relationship for tobacco products. Future studies are needed to understand factors that might contribute to this e-cigarette-specific path. Nonetheless, this path is much weaker compared to the general susceptibility-onset path. Taken as a whole, a general approach toward tobacco prevention supplemented with an e-cigarette-specific prevention may maximize the prevention of onset of tobacco use among youth in the US.

Findings from this study should be interpreted with the following limitations in mind. First, the study is observational in nature and does not provide definitive evidence for causal relationships. Second, we included a range of psychosocial variables in this study, but they are by no means exhaustive. Access to tobacco products, risk perceptions, use of other psychoactive drugs, maladaptive emotional and behavioral problems, family environment, neighborhood characteristics, and tobacco policies are all relevant predicts for tobacco use.^4,23-25^ As shown in our results, although approximately the same portions of youth were susceptible to cigarettes and e-cigarettes, the onset of e-cigarettes was much higher compared to the onset of cigarette smoking, which highlights the role of other variables in youth tobacco onset. Although out of the scope of the current study, future studies with the consideration of a range of variables would help identify the most important predictors for youth tobacco onset. Third, in this study, we studied the relationship linking susceptibility with the onset of use, the first major milestone of tobacco use. Future studies are needed to investigate whether susceptibility predicts continued use. Lastly, peer tobacco use was not assessed for a few tobacco product categories, which precluded the examination of the relationship between susceptibility and peer use for these tobacco products.

Counterbalancing strengths include (a) a national representative sample; (b) the use of ACASI, which helps reduce reporting bias; (c) a prospective design; (d) use of a structural equation modeling approach, which enables a more nuanced view of the relationship linking susceptibility with tobacco onset; and (e) a focus on youth never tobacco users, which provides evidence relevant for prevention programs.

Taken together, our findings suggest that youth tobacco prevention planning may benefit from a holistic approach toward youth risky behavior rather than a targeted approach at individual tobacco product categories.

## Data Availability

Data used in this study are publicly available and were downloaded on Dec. 13, 2019.

https://www.icpsr.umich.edu/icpsrweb/NAHDAP/studies/36498

https://www.icpsr.umich.edu/web/NAHDAP/studies/37786

## Authors’ contributions

Dr. Cheng has full access to all of the data in the study and takes responsibility for the integrity of the data and the accuracy of the data analysis.

Concept and design: Cheng, Largo, Knight.

Acquisition, analysis, or interpretation of data: Cheng, Lizhnyak.

Drafting of the manuscript: All authors.

Critical revision of the manuscript for important intellectual content: All authors.

Supervision: Vansickel, Largo

## Potential conflicts of interest

All authors are employees of Altria Client Services LLC.

---

**Supplementary Table 1.**
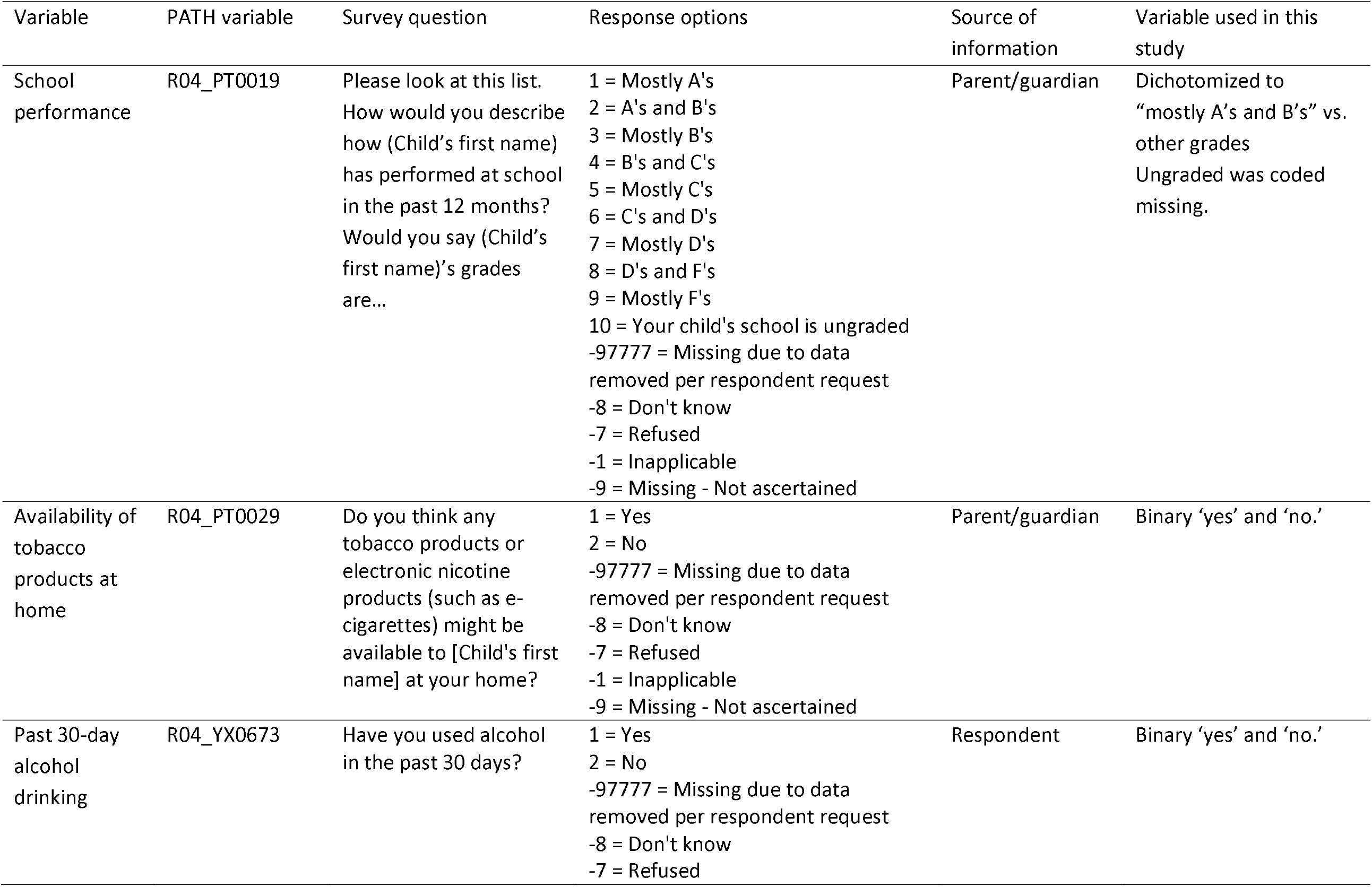
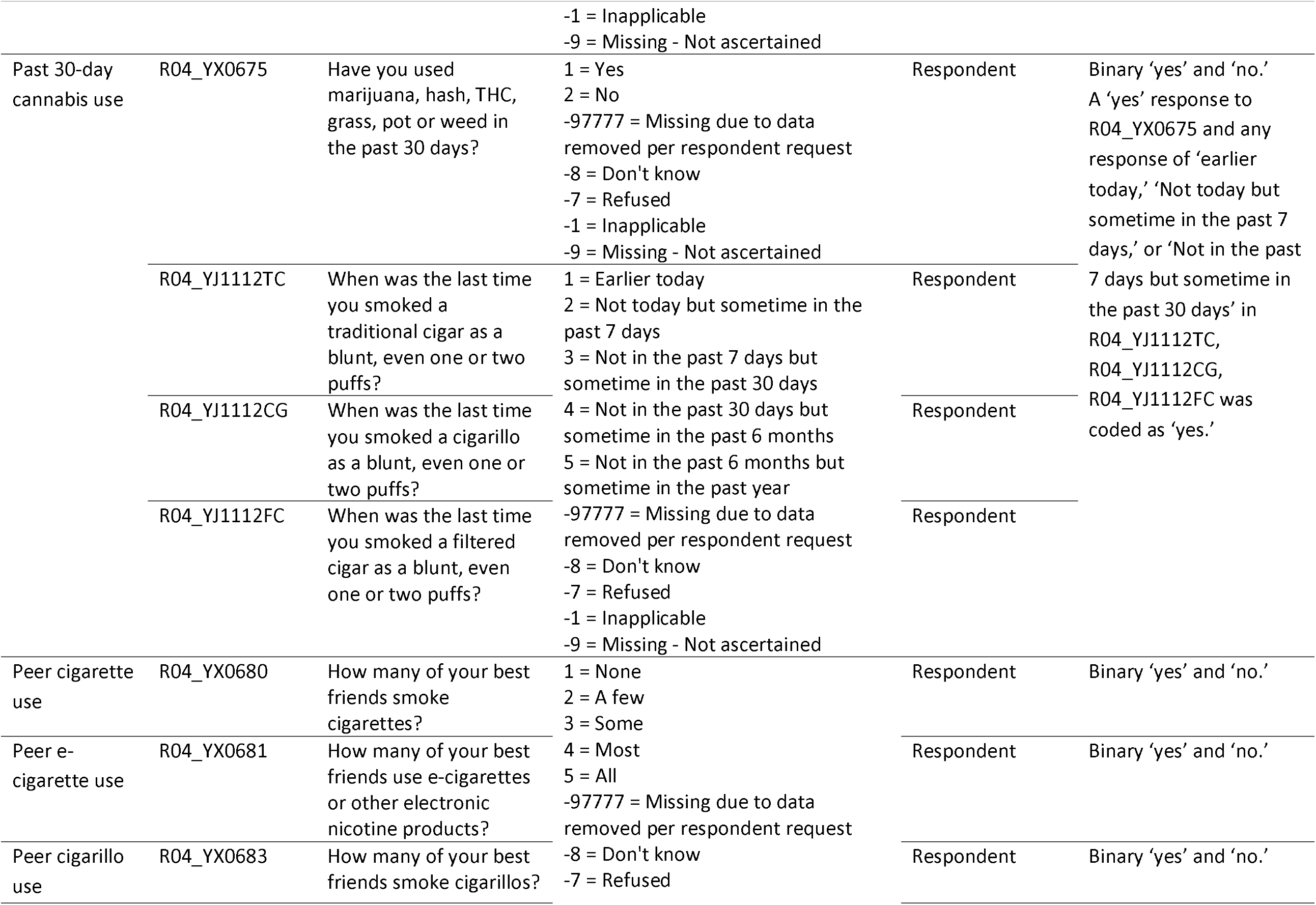
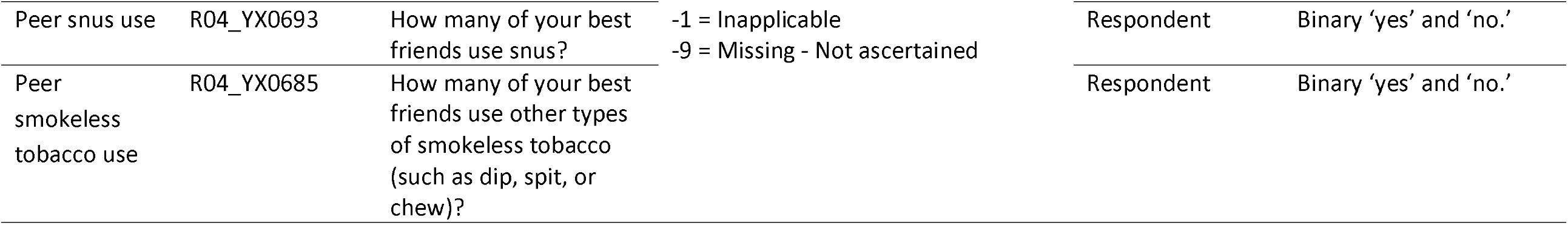
PATH wave 4 youth assessment of covariates used in this study.

## Notes

### Funding Statement

No external funding was received for this study.

### Author Declarations

The study involves analysis of publicly available data only. The original study was approved by an IRB.

## References

1. Pierce JP, Choi WS, Gilpin EA, Farkas AJ, Merritt RK. Validation of susceptibility as a predictor of which adolescents take up smoking in the United States. Health Psychol. 1996;15(5):355–361.

2. Gentzke AS, Creamer M, Cullen KA, et al. Vital Signs: Tobacco Product Use Among Middle and High School Students - United States, 2011-2018. MMWR Morb Mortal Wkly Rep. 2019;68(6):157–164.

3. Gentzke AS, Wang TW, Jamal A, et al. Tobacco Product Use Among Middle and High School Students - United States, 2020. MMWR Morb Mortal Wkly Rep. 2020;69(50):1881–1888.

4. Seo DC, Kwon E, Lee S, Seo J. Using susceptibility measures to prospectively predict ever use of electronic cigarettes among adolescents. Prev Med. 2020;130:105896.

5. Bold KW, Kong G, Cavallo DA, Camenga DR, Krishnan-Sarin S. E-Cigarette Susceptibility as a Predictor of Youth Initiation of E-Cigarettes. Nicotine Tob Res. 2018;20(4):527.

6. Cole AG, Kennedy RD, Chaurasia A, Leatherdale ST. Exploring the Predictive Validity of the Susceptibility to Smoking Construct for Tobacco Cigarettes, Alternative Tobacco Products, and E-Cigarettes. Nicotine Tob Res. 2019;21(3):323–330.

7. Chaffee BW, Cheng J. Tobacco product initiation is correlated with cross-product changes in tobacco harm perception and susceptibility: Longitudinal analysis of the Population Assessment of Tobacco and Health youth cohort. Prev Med. 2018;114:72–78.

8. Nicksic NE, Barnes AJ. Is susceptibility to E-cigarettes among youth associated with tobacco and other substance use behaviors one year later? Results from the PATH study. Prev Med. 2019;121:109–114.

9. Wang TW, Gentzke AS, Creamer MR, et al. Tobacco Product Use and Associated Factors Among Middle and High School Students - United States, 2019. MMWR Surveill Summ. 2019;68(12):1–22.

10. Cheng HG, Largo EG, Gogova M. E-cigarette use and onset of first cigarette smoking among adolescents: An empirical test of the ‘common liability’ theory. F1000Res. 2019;8:2099.

11. United States Department of Health and Human Services. Population Assessment of Tobacco and Health (PATH) Study [United States] Public-Use Files User Guide. In: Services DoHaH, ed. Ann Arbor, Michigan 48106: Inter-university Consortium for Political and Social Research; 2017.

12. Strong DR, Hartman SJ, Nodora J, et al. Predictive Validity of the Expanded Susceptibility to Smoke Index. Nicotine Tob Res. 2015;17(7):862–869.

13. Pierce JP, Distefan JM, Kaplan RM, Gilpin EA. The role of curiosity in smoking initiation. Addict Behav. 2005;30(4):685–696.

14. Steiger JH. Structural Model Evaluation and Modification: An Interval Estimation Approach. Multivariate Behav Res. 1990;25(2):173–180.

15. Bentler PM. Comparative fit indexes in structural models. Psychol Bull. 1990;107(2):238–246.

16. Hu Lt, Bentler PM. Cutoff criteria for fit indexes in covariance structure analysis: Conventional criteria versus new alternatives. Structural Equation Modeling: A Multidisciplinary Journal. 1999;6(1):1–55.

17. Bentler PM, Bonett DG. Significance tests and goodness of fit in the analysis of covariance structures. Psychological Bulletin. 1980;88(3):9.

18. Silveira ML, Conway KP, Everard CD, Sim HY, Kimmel HL, Compton WM. Longitudinal associations between susceptibility to tobacco use and the onset of other substances among U.S. youth. Prev Med. 2020;135:106074.

19. Vanyukov MM, Ridenour TA. Common liability to drug addictions: theory, research, practice. Drug Alcohol Depend. 2012;123 Suppl 1:S1–2.

20. Khouja JN, Wootton RE, Taylor AE, Davey Smith G, Munafo MR. Association of genetic liability to smoking initiation with e-cigarette use in young adults: A cohort study. PLoS Med. 2021;18(3):e1003555.

21. Farrelly MC, Duke JC, Nonnemaker J, et al. Association Between The Real Cost Media Campaign and Smoking Initiation Among Youths - United States, 2014-2016. MMWR Morb Mortal Wkly Rep. 2017;66(2):47–50.

22. Wills TA, Knight R, Williams RJ, Pagano I, Sargent JD. Risk factors for exclusive e-cigarette use and dual e-cigarette use and tobacco use in adolescents. Pediatrics. 2015;135(1):e43–51.

23. Resnick MD, Bearman PS, Blum RW, et al. Protecting adolescents from harm. Findings from the National Longitudinal Study on Adolescent Health. JAMA. 1997;278(10):823–832.

24. Fagan AA, Wright EM, Pinchevsky GM. The protective effects of neighborhood collective efficacy on adolescent substance use and violence following exposure to violence. J Youth Adolesc. 2014;43(9):1498–1512.

25. Aldrich MC, Hidalgo B, Widome R, Briss P, Brownson RC, Teutsch SM. The role of epidemiology in evidence-based policy making: a case study of tobacco use in youth. Ann Epidemiol. 2015;25(5):360–365.

